# Pregnancy episodes in All of Us: Harnessing multi-source data for pregnancy-related research

**DOI:** 10.1101/2024.04.10.24305609

**Authors:** Louisa H. Smith, Wanjiang Wang, Brianna Keefe-Oates

## Abstract

**Objective:** The National Institutes of Health’s All of Us Research Program addresses gaps in biomedical research by collecting health data from diverse populations. Pregnant individuals have historically been underrepresented in biomedical research, and pregnancy-related research is often limited by data availability, sample size, and inadequate representation of the diversity of pregnant people. We aimed to identify pregnancy episodes with high-quality electronic health record (EHR) data in All of Us Research Program data and evaluate the program’s utility for pregnancy-related research.

**Materials and Methods:** We used an algorithm to identify pregnancy episodes in All of Us EHR data. We described these pregnancies, validated them with additional data, and compared them to national statistics.

**Results:** Our study identified 18,970 pregnancy episodes from 14,234 participants; other possible pregnancy episodes had low-quality or insufficient data. Validation against people who reported a current pregnancy on an All of Us survey found low false positive and negative rates. Demographics were similar in some respects to national data; however, Asian-Americans were underrepresented, and older, highly educated pregnant people were overrepresented.

**Discussion:** Our approach demonstrates the capacity of All of Us to support pregnancy research and reveals the diversity of the pregnancy cohort. However, we noted an underrepresentation among some demographics. Other limitations include measurement error in gestational age and limited data on non-live births.

**Conclusion:** The wide variety of data in the All of Us program, encompassing EHR, survey, genomic, and Fitbit data, offers a valuable resource for studying pregnancy, yet care must be taken to avoid biases.

## INTRODUCTION

Despite the critical role of pregnancy in human health and development, it remains an understudied area in biomedical research. The complexities and ethical considerations of studying the pregnant population have led to the widespread exclusion of pregnant people from clinical trials and efficacy studies, making observational data necessary to study the impacts of various exposures, as well as to gain insights into the broader spectrum of pregnancy-related health conditions, behaviors, and outcomes. Nevertheless, research into pregnancy and the postpartum period remains challenging due to data and study design limitations.

Historically, studies about pregnancy and postpartum health outcomes have relied on costly cohort studies or surveillance mechanisms that do not capture the entire pregnancy period. For example, state and national governments provide representative but cross-sectional birth surveillance data that also fails to capture early outcomes or sufficient data on pregnancy exposures such as medications.^1,2^ Birth cohorts, observational studies that recruit pregnant or recently postpartum people and their infants, often recruit participants in the later stages of pregnancy, missing the earliest pregnancy exposures and outcomes such as miscarriages.^3–5^ More recently, preconception cohort studies have been designed to prospectively collect data on fertility, pregnancy, and postpartum health outcomes.^6–9^ While cohort studies are rich in longitudinal data, they are also inherently resource-intensive and often rely primarily on self-report of pregnancy timing and outcomes. Real-world data, including electronic health records (EHR) and insurance claims, contain thorough information about diagnoses, medications, and healthcare procedures and their costs. While the longitudinal nature of the data, coupled with very large sample sizes, is promising for studying key biomedical outcomes, these datasets usually lack important demographic information.^10^ Additionally, identifying pregnancies and measuring gestational age in these complicated records is challenging.

The All of Us Research Program provides an opportunity for pregnancy-related research that overcomes some of the limitations of other data sources. The program collects information through EHR and survey data, as well as biospecimens and data from activity trackers, allowing for longitudinal studies examining both social and biomedical exposures and outcomes. Conducting pregnancy-related research using All of Us Research Program data aligns well with the program’s mission, which is to fully represent the diversity of the US population by explicitly including groups historically underrepresented in biomedical research.^11^ Pregnant people, and in particular pregnant people of color and sexual and gender minorities,^12–14^ make up an important and understudied segment of this population.

While this research program provides an opportunity to ask new research questions about pregnancy, challenges to identifying and characterizing pregnancies in real-world data remain. Identifying pregnancy episodes in the All of Us EHR data can be difficult due to variations in coding practices, patients visiting multiple healthcare providers over the course of a pregnancy, and infrequent use of codes that identify gestational age. These challenges mean that researchers can’t rely on a single code to identify pregnancy, and complex algorithms are needed to identify pregnancies, ascertain their outcomes, and estimate gestational ages.^15^

## OBJECTIVE

This work aimed to identify pregnancy episodes in the All of Us population. Objectives included inferring gestational age and pregnancy outcomes, validating episodes using survey data, and characterizing the identified pregnancies. We aimed to establish how the All of Us Research Program could be used to answer pregnancy-related research questions important to researchers and the communities they serve.

## METHODS

### Data

The All of Us Research Program began enrolling adult participants across the country in 2018.^11^ Focused recruitment occurs at an extensive network of sites nationwide; the study is also open to any volunteer. Participants must complete a baseline survey with demographic information; additional surveys collect data on health history, social context, and more. Volunteers may also link EHR data and contribute biospecimens for genomic analyses; movement, heart rate, and sleep data through their Fitbit device; clinic-based body measurements, and more. Due to the geographic diversity of the participants, EHR data is contributed by a large number of institutions with different medical record systems. The transformation of the data into the Observational Medical Outcomes Partnership common data model (OMOP CDM)^16,17^ allows the EHR data to be combined and analyzed across participants on a web-based platform. After applying quality control and privacy-preserving measures, data is released to researchers who complete the required training and data use agreement. This study used data from the *All of Us* Research Program’s

Controlled Tier Dataset Version 7 (release C2022Q4R9), available to authorized users on the Researcher Workbench. The project followed the guidelines for ethical conduct of research put in place by All of Us and was determined to be exempt by the Northeastern IRB.

### Pregnancy identification algorithm

We identified pregnancy episodes among All of Us participants contributing EHR data using an algorithm developed and validated in the National Covid Cohort Collaborative data.^15^ This approach, Hierarchy and rule-based pregnancy episode Inference integrated with Pregnancy Progression Signatures [HIPPS], consists of three sub-algorithms: Hierarchy-based Inference of Pregnancy (HIP), Pregnancy Progression Signature (PPS), and Estimated Start Date (ESD). These are described here briefly; a complete description is available in the original publication.^15^

The HIP algorithm defines pregnancy episodes based on how a pregnancy ended: live birth, stillbirth, ectopic pregnancy, spontaneous or induced abortion, and delivery (delivery-only codes are non-specific to the outcome of the pregnancy). First, OMOP concept codes related to these outcomes are identified and then deduplicated within patients based on minimum plausible pregnancy durations (e.g., 182 days between delivery outcomes and 56 days between consecutive ectopic pregnancies). These outcome-based episodes are combined with overlapping gestation-based episodes, which are defined by the timing of a participant’s minimum and maximum gestational-age-related codes (e.g., “Gestation period, 36 weeks”).

The PPS algorithm relies on a different but overlapping set of initial pregnancy-related codes. (We used the codes identified by Jones et al.^15^ rather than identifying codes specific to All of Us, due to the similarity of the data sources.) Along with codes for individual gestational weeks, PPS uses codes specific to a broader gestational age range (e.g., glucose tolerance tests most often occur within 6 and 8 months of gestation). The algorithm identifies episodes with plausible progressions of these timing-specific concepts. Next, pregnancy outcome codes that overlap with the expected timing relative to the gestational age codes are used to assign outcomes to the episodes.

The HIP-identified and PPS-identified episodes are then merged based on overlapping timing. The ESD algorithm uses the gestational age codes to assign an inferred pregnancy start date and a date the pregnancy outcome occurred, along with the corresponding gestational age. Both week-specific and gestational-age range codes are used to identify and remove outlying timing codes; the inferred start and end dates are based on the latest and most specific codes.

As in Jones et al.,^15^ we classified the pregnancy episodes based on whether the pregnancy outcomes from both the HIP and PPS algorithms matched, the dates of those outcomes were within 14 days of each other, and whether the estimated gestational age at the end of pregnancy was plausible. We assigned episodes a concordance score of 2 if all three criteria were met, a 1 if the outcome didn’t match, and a 0 otherwise. We also classified episodes based on how specific the gestational age concepts were: “non-specific” if there were no gestational age-related codes or if the timing could not be narrowed down below a three-month window, “1-3 months” or “1-3 weeks” if gestational age could be narrowed to one of those windows, and “1 week (poor support)” if there was only a single gestational week concept within a pregnancy episode.

We translated the original code for the algorithm from PySpark SQL to work in R on the Researcher Workbench, where we primarily used dbplyr^18^ and the allofus R package.^19^ All of our analysis code is available on Github (https://github.com/louisahsmith/allofus-pregnancy).

### Validation

We validated the pregnancy episodes in two ways: with survey data and with pregnancy-related codes that were not used in the HIPPS algorithm. First, we identified All of Us participants who responded to the Overall Health survey, contributed EHR data, were aged 15-55, and didn’t report male sex at birth. We compared those participants’ responses to the question “Are you currently pregnant?” and the identified pregnancy episodes to assess misclassification. We considered a false positive pregnancy episode to be one in which a participant reported not being pregnant on a date they were identified as greater than 12 weeks pregnant by the algorithm, and a missed episode (false negative) to occur when someone reported being pregnant at a time not overlapping with an identified pregnancy episode (Figure 1). Second, we quantified occurrences of 25 pregnancy-related codes not used in identifying the pregnancies among all participants and compared their frequency during the expected pregnancy timing to the overall population frequency.

**Figure 1.**
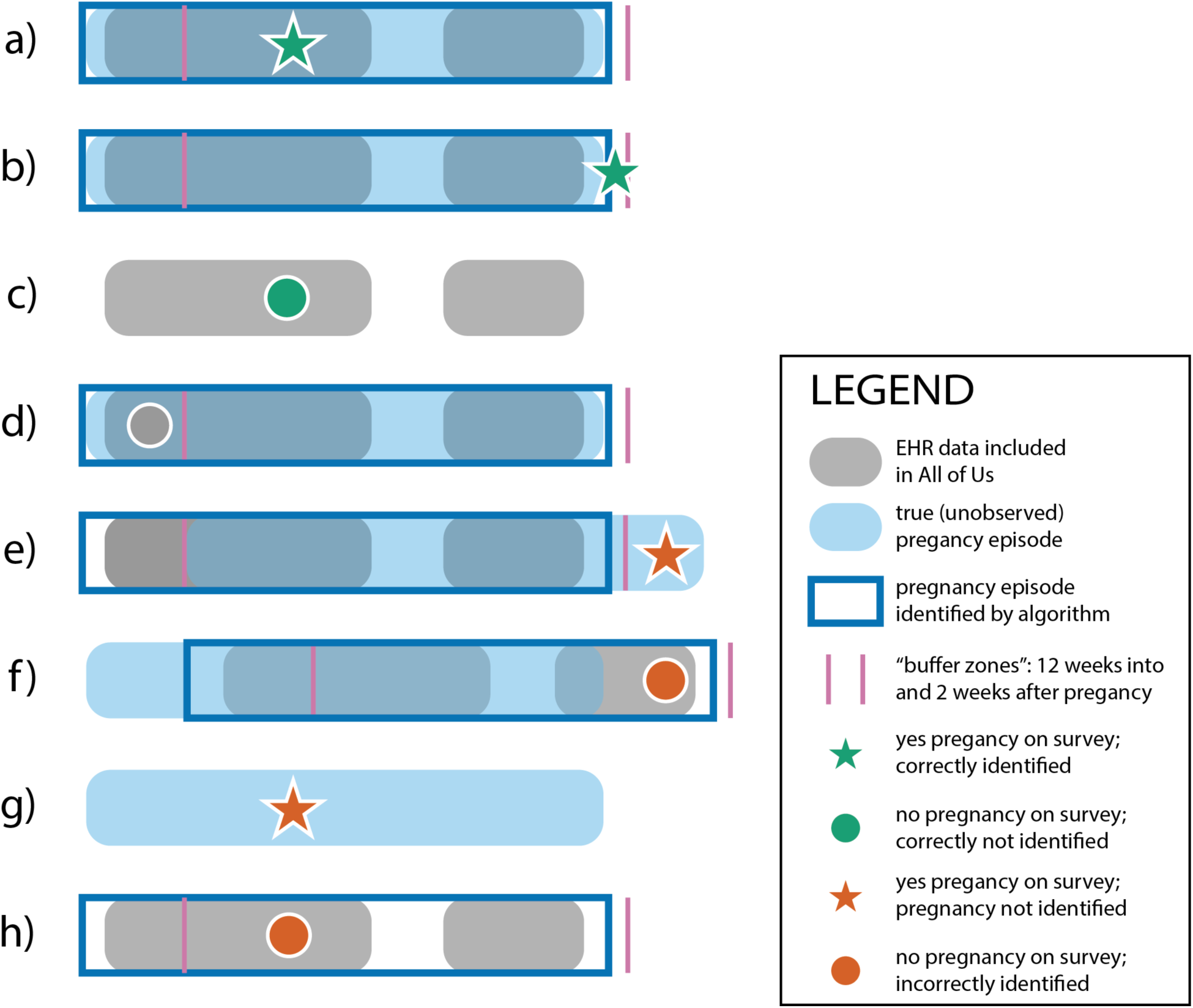
Schematic describing the validation study using survey data and scenarios resulting in errors. The participants represented in a) and b) reported being pregnant during, or within two weeks of, an assigned pregnancy episode, representing true positives. Participant c) reported not being pregnant during a time they were not assigned a pregnancy episode, representing a true negative. Participant d) answered the survey before 12 weeks of an assigned pregnancy episode, meaning we didn’t know whether they would have known they were pregnant at the time (these situations were excluded from the validation). Participant e) represents a situation in which the identified pregnancy episode is misaligned with the truth, resulting in a false negative. The setting of participant f) was an apparently more common form of misalignment, in which the assigned pregnancy episode presumably was delayed relative to the truth, representing a false positive. The situation in g) was also a common error, in which there was no or very little EHR data available around the time of a positive response to the survey question (i.e., a false negative for the algorithm), suggesting that the data containing pregnancy information was not included in the All of Us database. Participant h) has a false positive identified pregnancy episode not closely linked in time with any true pregnancies, a situation that occurred when pregnancy-related codes were apparently carried forward for years after their first occurrence.

### Characteristics of pregnancies and pregnant All of Us participants

We characterized the pregnancy episodes by their outcomes, gestational lengths, year of pregnancy, and demographic characteristics of the pregnant people. We fit exploratory log-linear regression models to describe characteristics associated with a higher probability of having multiple pregnancy episodes, having a live birth vs. another pregnancy outcome, and delivering preterm (among live births). We also characterized the extent to which pregnant participants contributed additional types of data to All of Us, including survey, Fitbit, and genomic data.

We used public vital statistics data to compare the demographics of All of Us pregnancies ending in live births to national statistics.^20^ Specifically, we computed age, education, racial/ethnic, and state breakdowns of US live births from 2016-2022 and standardized those to the distribution of pregnancies by calendar year in All of Us.

## RESULTS

### Pregnancy episodes

There were 134,566 individuals in the Controlled Tier C2022Q4R9 release of All of Us (participant data cutoff date of 7/1/2022) who did not report male sex at birth and who had contributed EHR data at some point between the ages of 15 and 55. Duration of retrospective EHR data varies by participant and contributing data site; we used data from as early as 1979. Overall, we identified 59,646 pregnancy episodes among 31,726 unique All of Us participants. Of these episodes 30,177 were identified by both the HIP and PPS algorithms, and 31,518 occurred since 2016. Concordance differed over time, with earlier pregnancies less likely to be identified by both algorithms or result in matching outcomes and dates (Figure 2). Among pregnancies since 2016 identified by both algorithms (n = 18,970), concordance was high, with 83.4% (n = 15,822) fully concordant and an additional 9.6% (n = 1,815) with concordant dates and plausible gestational age but mismatched outcomes (e.g., live birth vs. delivery). Due to the unreliability of early EHR data, we focus our main results on the pregnancy episodes occurring 2016-2022 that were identified by both algorithms (“high-quality episodes”) except when otherwise specified.

**Figure 2.**
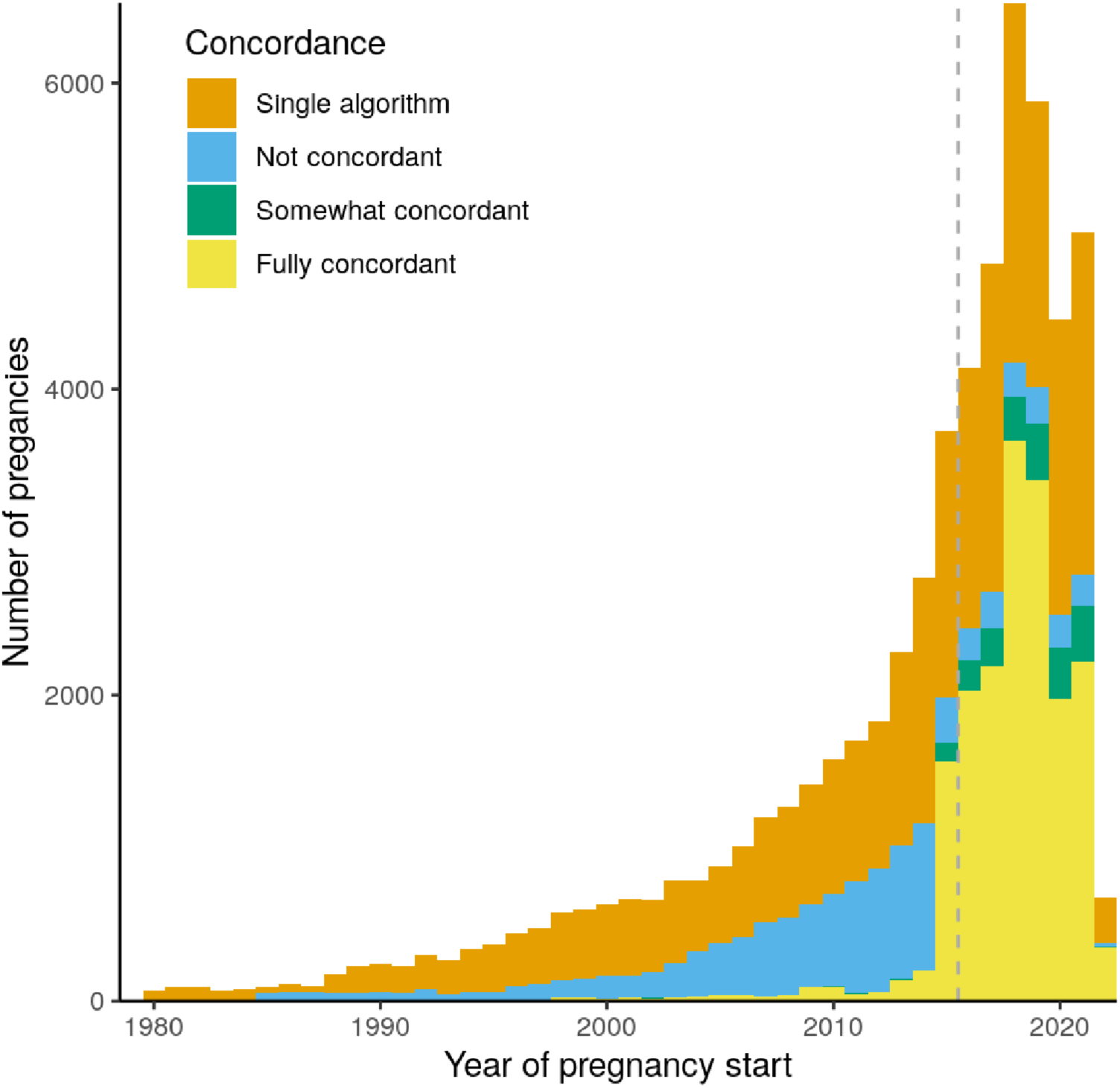
Concordance between pregnancy identification algorithms by date across all pregnancies. Fully concordant pregnancies have matching HIP and PPS outcomes, similar end dates (within 14 days), and a plausible gestational age. Somewhat and not concordant episodes differed on outcome category or timing. Additional episodes were identified by only one of the two algorithms. We included episodes starting after 2016 (dashed line) that were identified by both algorithms in our main analysis.

The majority of the high-quality pregnancy episodes ended in live birth (n = 11,379; 60.0%); 26.7% (n = 5,053) were missing an outcome (Figure 3). Gestational age was assigned to a specific week for 77.9% (n = 14,785) of pregnancy episodes; just 3.8% (n = 727) had non-specific gestational duration information (Supplementary Table 1).

**Figure 3.**
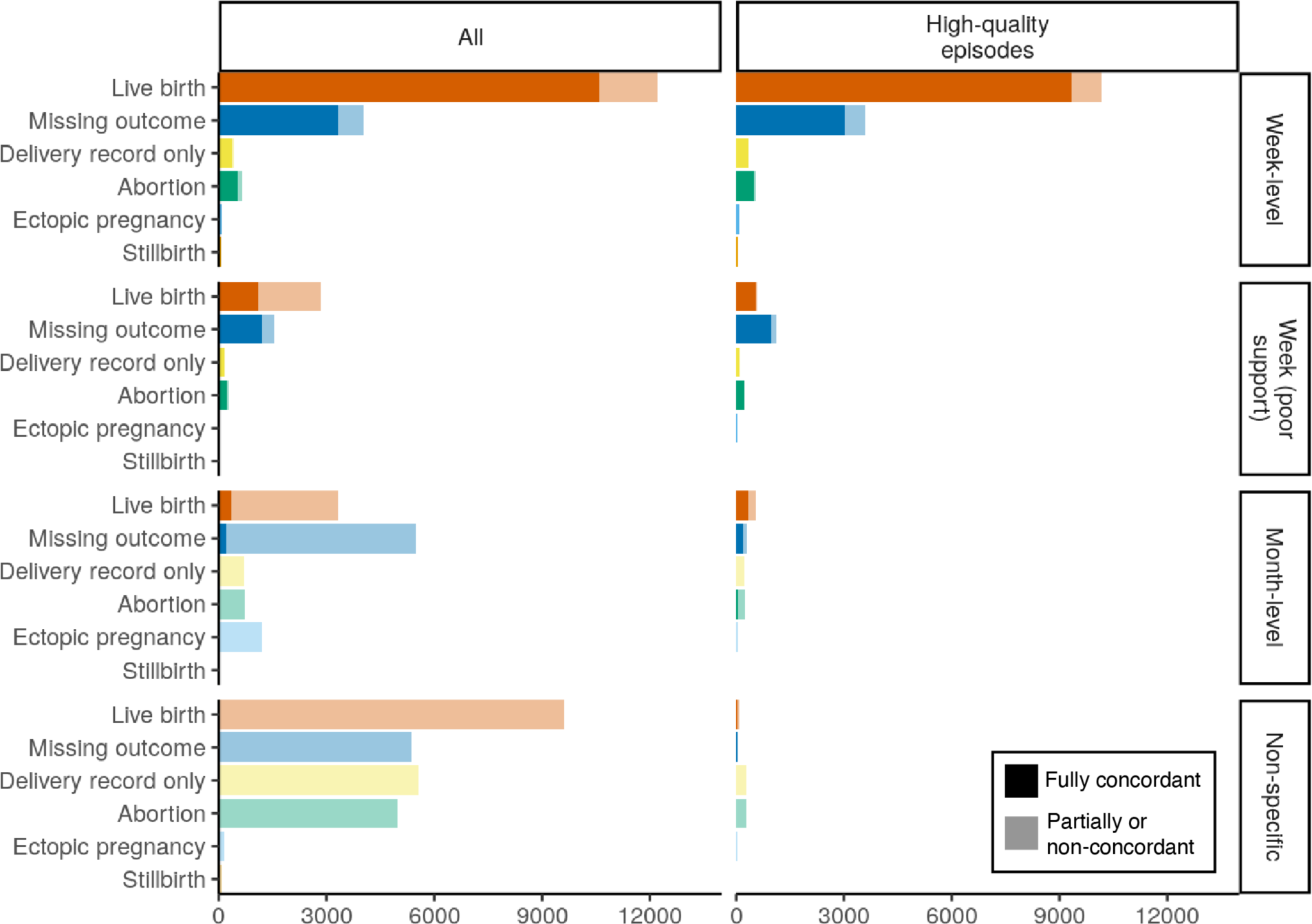
Distribution of pregnancy outcomes among all identified pregnancies (left panel) and the high-quality episodes (right panel). Episodes are stratified by the precision of the gestational age information used to assign pregnancy timing. Week-level pregnancy episodes were able to be dated to within less than a month; those with poor support only had a single week-specific code. Month-level episodes were dated to within 1-3 months, and non-specific episodes were less precise. In addition, shaded portions of the bars represent the highly concordant episodes, on which both HIP and PPS algorithms agreed on outcome and timing, and light-colored portions represent partial or non-concordance.

### Validation

There were few false positives and false negatives as determined by the self-reported pregnancy status survey question. There were 63,419 All of Us participants who answered “yes” (n = 4,680) or “no” (n = 58,739) to the pregnancy question on the “Overall Health” survey whose potential pregnancies we could capture in EHR data. Of those reporting current pregnancy, 3,832 (sensitivity = 81.8%) had an identified pregnancy episode within 2 weeks of the survey date. Compared to the 848 survey respondents reporting a pregnancy we didn’t identify as overlapping, those we captured had more EHR data both overall (mean 90.1 codes vs. 35.9 codes) and specific to pregnancy (mean 12.0 codes vs. 2.3 codes of those used in HIP algorithm). In addition, more of their EHR codes occurred post-survey (mean time post-survey 62 days vs. -7 days), suggesting that their pregnancies were not identified because EHR data containing their pregnancy outcomes had not (yet) been added to the All of Us dataset (Supplementary Table 2).

Specificity was over 99%, with 518 respondents reporting no pregnancy despite our algorithm identifying them as more than 12 weeks pregnant. Of these, the median time from the survey date to the identified pregnancy end date was 14 days (interquartile range 0, 19.8 days), suggesting that these participants took the survey soon after pregnancy, but the dates related to the pregnancy outcome codes in their EHR data were delayed. The positive predictive value was 83.1%, and the negative predictive value was 98.5%.

Overlap of the 25 clinician-curated concepts not used in the HIPPS algorithm to identify pregnancies varied, in part because we only considered the high-quality episodes to be matches. Overall, 63.8% of occurrences were during an identified episode at an appropriate gestational age, though some overlapped substantially less (e.g., 28.1% of occurrences of genetic counseling services; Supplementary Table 3).

### All of Us participant characteristics

Of the 14,234 All of Us participants with at least one high-quality pregnancy episode, most were Hispanic or Latino (43.2%) or non-Hispanic White (33.6%) (Table 1). Pregnant people represented 41 US states and territories, but over half had fewer than 10 participants; over two-thirds were from just four states: Arizona, New York, California, and Massachusetts. The vast majority reported being women (99.4%) and heterosexual (91.8%).

**Table 1.**
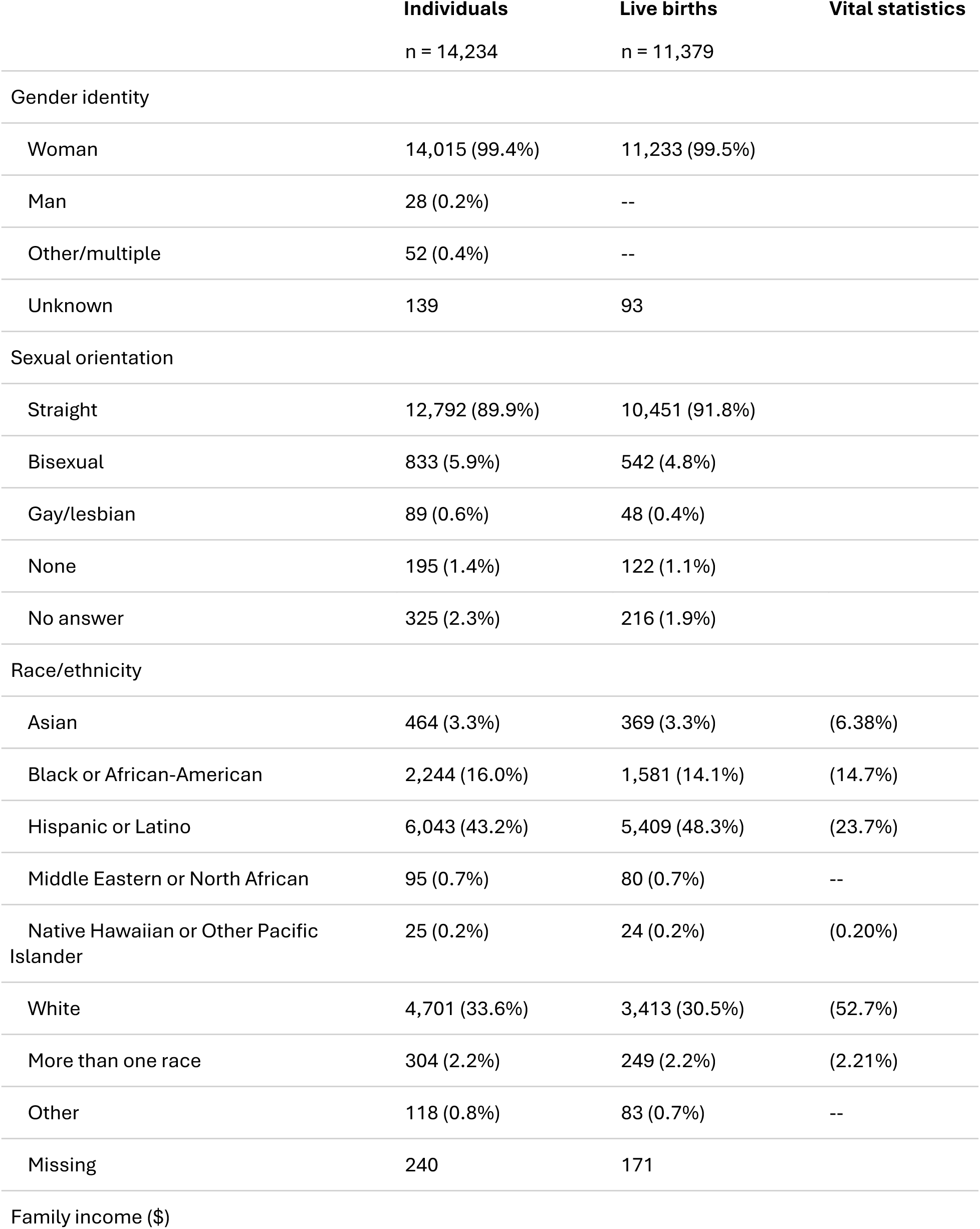
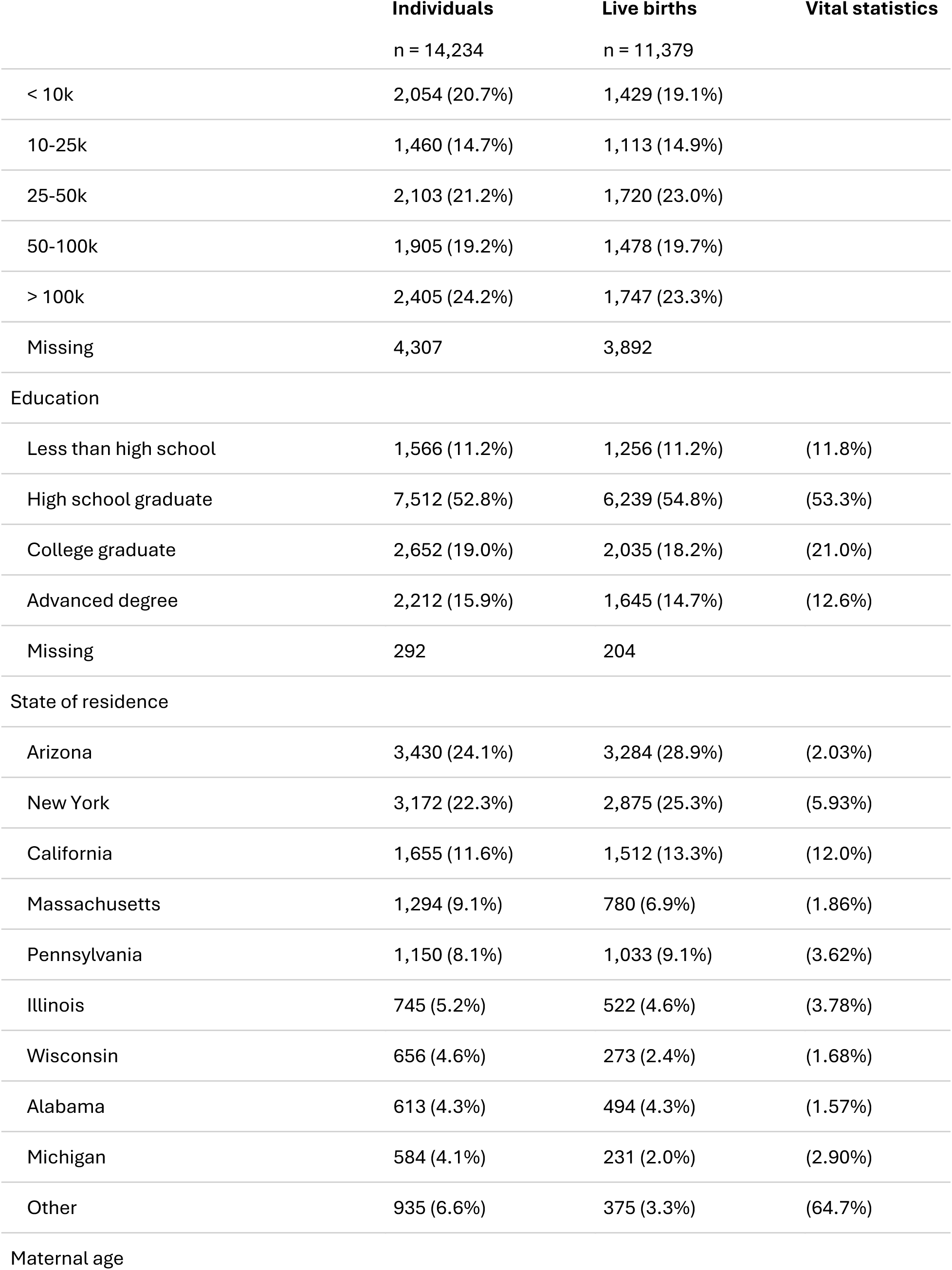
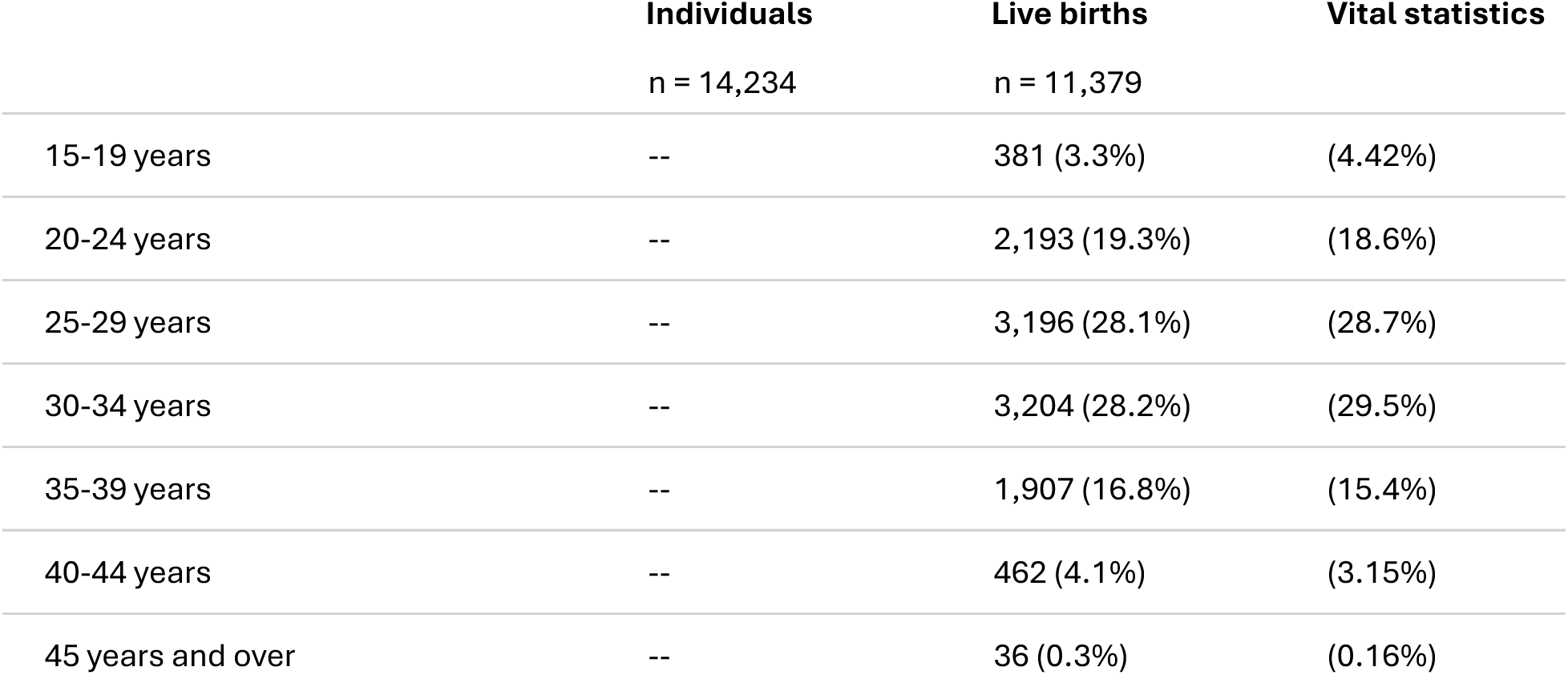
Demographic characteristics of All of Us participants with high-quality pregnancy episodes and of live births, compared to vital statistics. Live births may represent multiple pregnancies from the same participant. Vital statistics data have been standardized to the distribution of delivery years in the All of Us data.

|  | **Individuals**<br>n = 14,234 | **Live births**<br>n = 11,379 | **Vital statistics** |
| --- | --- | --- | --- |
| **Gender identity** |  |  |  |
| Woman | 14,015 (99.4%) | 11,233 (99.5%) |  |
| Man | 28 (0.2%) | -- |  |
| Other/multiple | 52 (0.4%) | -- |  |
| Unknown | 139 | 93 |  |
| **Sexual orientation** |  |  |  |
| Straight | 12,792 (89.9%) | 10,451 (91.8%) |  |
| Bisexual | 833 (5.9%) | 542 (4.8%) |  |
| Gay/lesbian | 89 (0.6%) | 48 (0.4%) |  |
| None | 195 (1.4%) | 122 (1.1%) |  |
| No answer | 325 (2.3%) | 216 (1.9%) |  |
| **Race/ethnicity** |  |  |  |
| Asian | 464 (3.3%) | 369 (3.3%) | (6.38%) |
| Black or African-American | 2,244 (16.0%) | 1,581 (14.1%) | (14.7%) |
| Hispanic or Latino | 6,043 (43.2%) | 5,409 (48.3%) | (23.7%) |
| Middle Eastern or North African | 95 (0.7%) | 80 (0.7%) | -- |
| Native Hawaiian or Other Pacific Islander | 25 (0.2%) | 24 (0.2%) | (0.20%) |
| White | 4,701 (33.6%) | 3,413 (30.5%) | (52.7%) |
| More than one race | 304 (2.2%) | 249 (2.2%) | (2.21%) |
| Other | 118 (0.8%) | 83 (0.7%) | -- |
| Missing | 240 | 171 |  |
| **Family income (\$)** | | | |

|  | Individuals | Live births | Vital statistics |
| --- | --- | --- | --- |
|  | n = 14,234 | n = 11,379 |  |
| < 10k | 2,054 (20.7%) | 1,429 (19.1%) |  |
| 10-25k | 1,460 (14.7%) | 1,113 (14.9%) |  |
| 25-50k | 2,103 (21.2%) | 1,720 (23.0%) |  |
| 50-100k | 1,905 (19.2%) | 1,478 (19.7%) |  |
| > 100k | 2,405 (24.2%) | 1,747 (23.3%) |  |
| Missing | 4,307 | 3,892 |  |
| Education |  |  |  |
| Less than high school | 1,566 (11.2%) | 1,256 (11.2%) | (11.8%) |
| High school graduate | 7,512 (52.8%) | 6,239 (54.8%) | (53.3%) |
| College graduate | 2,652 (19.0%) | 2,035 (18.2%) | (21.0%) |
| Advanced degree | 2,212 (15.9%) | 1,645 (14.7%) | (12.6%) |
| Missing | 292 | 204 |  |
| State of residence |  |  |  |
| Arizona | 3,430 (24.1%) | 3,284 (28.9%) | (2.03%) |
| New York | 3,172 (22.3%) | 2,875 (25.3%) | (5.93%) |
| California | 1,655 (11.6%) | 1,512 (13.3%) | (12.0%) |
| Massachusetts | 1,294 (9.1%) | 780 (6.9%) | (1.86%) |
| Pennsylvania | 1,150 (8.1%) | 1,033 (9.1%) | (3.62%) |
| Illinois | 745 (5.2%) | 522 (4.6%) | (3.78%) |
| Wisconsin | 656 (4.6%) | 273 (2.4%) | (1.68%) |
| Alabama | 613 (4.3%) | 494 (4.3%) | (1.57%) |
| Michigan | 584 (4.1%) | 231 (2.0%) | (2.90%) |
| Other | 935 (6.6%) | 375 (3.3%) | (64.7%) |
| Maternal age |  |  |  |
| 15-19 years | -- | 381 (3.3%) | (4.42%) |
| 20-24 years | -- | 2,193 (19.3%) | (18.6%) |
| 25-29 years | -- | 3,196 (28.1%) | (28.7%) |
| 30-34 years | -- | 3,204 (28.2%) | (29.5%) |
| 35-39 years | -- | 1,907 (16.8%) | (15.4%) |
| 40-44 years | -- | 462 (4.1%) | (3.15%) |
| 45 years and over | -- | 36 (0.3%) | (0.16%) |

People with incomes greater than $100,000 per year were more likely than any other income bracket to have more than one pregnancy episode captured, as were those who were married/partnered or divorced/separated/widowed compared to never married (Supplementary Table 4). There was a steep decline with age in the probability that a pregnancy episode ended in a live birth, with risk ratios of 0.93 (95 % CI 0.87, 1.00) at 35-34 years and 0.63 (95 % CI 0.41, 0.91) at 45-49 years compared to 25-29 years (Supplementary Table 5). Black participants were more likely than other race/ethnic groups to have preterm deliveries, as were older compared to younger participants (Supplementary Table 6).

Almost all participants with high-quality pregnancy episodes completed the Lifestyle and Overall Health surveys along with the required Basics survey (Table 2). In addition, 36.1% completed the Personal and Family Medical survey, 35.3% the Healthcare Access survey, and 14.5% the Social Determinants of Health survey. Few have contributed Fitbit data during their pregnancy (n = 211 with activity data; n = 176 with heart rate data; n = 195 with sleep data), but 88.9% have some genomic data available. Most pregnancy episodes (49.2%) occurred before joining All of Us, but a substantial number joined during pregnancy (24.4%) or had prospective pregnancy episodes (26.2%).

**Table 2.** Additional All of Us data contributed by participants with identified high-quality pregnancy episodes.

|  |  |
| --- | --- |
| **All of Us participation relative to pregnancy episode**<br>(n = 18,970 episodes) |  |
| After | 9364 (49.4%) |
| Before | 5079 (26.8%) |
| During | 4459 (23.5%) |
| Unclear | 68 (0.4%) |
| **Fitbit device data during pregnancy episode**<br>(n = 18,970 episodes) |  |
| Activity | 211 (1.1%) |
| Sleep | 195 (1.0%) |
| Heart Rate | 176 (0.9%) |
| **Genomic data**<br>(n = 14,234 individuals) |  |
| Array data | 12,650 (88.9%) |
| Whole genome variant data | 10,259 (72.1%) |
| Long read whole genome variant | 48 (0.3%) |
| Structural variant | 506 (3.6%) |
| **Survey data**<br>(n = 14,234 individuals) |  |
| The Basics | 14,234 (100%) |
| Lifestyle | 14,230 (100%) |
| Overall Health | 14,230 (100%) |
| Personal/Family Health History | 5,139 (36.1%) |
| Healthcare Access and Utilization | 5,021 (35.3%) |
| Social Determinants of Health | 2,071 (14.5%) |

### Live births

Among live births, the median gestational age was 39.0 (interquartile range 37.1, 39.0), and 20.5% were inferred to have delivered before 37 weeks gestation (i.e., preterm). Compared to vital statistics data from the same years (Table 1), live births in All of Us were to slightly older (21.1% vs. 18.7% 35 years or greater) and more educated (14.5% vs. 12.6% with a graduate degree) individuals. Similar proportions of births were to Black, Native Hawaiian/Pacific Islander, or individuals with more than one race. However, compared to national data, All of Us had a smaller proportion of Asian (3.3% vs. 6.4%) and non-Hispanic White (30.9% vs 52.7%), and more Hispanic or Latino (49.0% vs. 23.7%) mothers. Vital statistics do not capture Middle Eastern/North African ethnicity, but 95 All of Us participants reporting that ethnicity had high-quality pregnancy episodes (Table 1).

## DISCUSSION

In this study, we used All of Us EHR data to identify pregnancy episodes and estimate gestational age. In doing so, we validated an algorithm recently developed for use in OMOP CDM data and demonstrated the capability of the All of Us data to support pregnancy research with a diverse cohort of pregnant people.

### Promise and potential of All of Us multi-source data

Maternal morbidity and mortality are of utmost concern in the US, with rising risks and severe disparities in outcomes by race and ethnicity.^21,22^ The ability to link health outcome data from medical records to survey questions about health behaviors, medical history, and lifestyle and social factors can provide new insights into experiences during pregnancy and in the postpartum period. In particular, the survey data collected by All of Us provides in-depth information about social determinants of health that may help explain disparities in maternal health and, most importantly, identify interventions. The over 4000 people who apparently completed All of Us surveys while pregnant make for a significant cohort that can be followed through the postpartum period, allowing researchers to ask questions about relationships between social factors such as social support and experiences of discrimination in pregnancy and postpartum health and well-being.

Other sources of data, including genomic and activity device data, provide additional opportunities to answer critical pregnancy-related questions. For example, the causes of preterm delivery appear to be numerous but are poorly understood;^23,24^ linking the inferred gestational ages of the All of Us pregnancies with these sources of big data might produce new insights. Future data types on the All of Us data roadmap^25^ include self-reported height and weight, activity tracker data from Apple’s popular platform, and data from a nutrition substudy, all of which could provide more data to study predictors and outcomes associated with weight and nutrition during preconception, pregnancy, and postpartum.

Activity tracker data shows promise not only for research on physiologic changes during pregnancy but also for inclusion in an improved pregnancy identification algorithm. Epstein and McCoy used EHR and Fitbit data in 89 All of Us participants to observe the change in heart rate for pregnant people and found a peak in heart rate during the first and third trimesters and a steady increase through the second trimester.^26^ Other research also showed that integrating EHR with wearable data can be used in predictive models for hospital readmission.^27^ Given potential variations in daily exercise, heart rate, and sleep duration, integrating data from wearable devices could augment the algorithm’s effectiveness.

### Strengths and limitations of EHR pregnancy data

Real-world data like the EHR data used in this study offers valuable insights into pregnancies and health-related characteristics of pregnant people. Additionally, it spans many years of patient health records, enabling a comprehensive understanding of medical history, including pre-pregnancy and post-pregnancy phases, and providing a holistic view of participant health. Furthermore, compared to clinical trial data, which generally has stringent inclusion/exclusion criteria, EHR data like that in All of Us better represent real-world populations, including underrepresented groups, fostering a more inclusive, reliable, and comprehensive approach to research.^28^

However, EHR data comes with its own limitations. Firstly, coding errors lead to incomplete or inaccurate documentation of patient records. Certain medical conditions may not be fully captured or explained by these codes, leaving important information conveyed through free text, which is not included in All of Us data. Secondly, patients often receive care from multiple healthcare systems, resulting in fragmented records that result in missing information. For instance, a patient receiving prenatal care at one hospital may need emergency labor services at a different hospital within another health system. Indeed, over one-quarter of the otherwise high-quality pregnancy episodes we identified were missing an outcome, though in some cases this was likely due to pregnancies that continued past the data cut-off point.

In our survey-based validation substudy that included people who joined All of Us while pregnant, we estimated sensitivity exceeding 80% and specificity approaching 100%, affirming the Jones et al approach for reliably identifying pregnancy episodes. Other studies have also reported high agreement rates and positive predictive values in line with our study, from 70% to close to 100%.^15,29–33^ However, we could not specifically validate gestational age at the outcome, which may be less accurately identified than the outcome itself, particularly for non-live birth outcomes.^34^ In addition, our approach to validation over-represented EHR information occurring around the time participants joined All of Us and took the surveys, when their healthcare is more likely to be occurring within systems that contribute data to All of Us, leading to an overestimate of the likely sensitivity of the algorithm over the entire scope of the data.

As with other pregnancy algorithms, HIPPS leverages medical codes that represent key factors such as prenatal care procedures, gestational age, and a range of pregnancy outcomes. The algorithm was developed for data that has been translated to the OMOP CDM, which is made up of a common vocabulary of concept codes representing other code sets, including ICD-9, ICD-10, and CPT codes. This makes the algorithm highly transferable to different settings and across time. Indeed, we identified pregnancies as early as the 1980s despite changes in medical coding since then. However, the early episodes were of notably lower quality based on concordance between algorithms and estimated gestational age, reflecting improvements in electronic health record keeping and the usage of gestational age-specific ICD-10 codes.^35,36^

### Recommendations and future directions

Properly accounting for timing is critical in pregnancy-related research and will be even more so in All of Us studies, as different data components are contributed at different times relative to a given pregnancy. Half of the pregnancies we identified occurred before a participant joined All of Us, limiting the sample size for research questions in which exposures of interest and covariates are drawn from survey questions. Researchers should be careful not to make analyses conditional on joining All of Us post-pregnancy; for example, pre-All of Us pregnancies are guaranteed not to have resulted in maternal mortality. Nonetheless, some of the survey responses (e.g., race/ethnicity) can be combined with EHR regardless of timing, as can genetic data. As All of Us grows, we can expect more prospective pregnancies to occur.

While All of Us aims to be inclusive, it is not necessarily representative. We found that in several respects, demographic data on pregnancies in All of Us did not match that from vital statistics. While not inherently a problem, researchers should consider selection as a source of bias in their studies, thinking carefully about who is joining All of Us and contributing each type of data. A lack of geographic diversity due to intense recruitment at some All of Us sites suggests that a lack of diversity is likely in other, unmeasured respects. In future work, we will consider how to address possible biases due to selection and missing data and improve the generalizability of the data.

Although the algorithm we used was not perfectly accurate even according to our limited validation exercises, the use of an algorithm like this one represents an improvement compared to a simple code search for pregnancy or delivery-related codes. While live birth is a relatively straightforward outcome to recognize, other outcomes, such as ectopic pregnancy, require more supporting information before a single code should be considered indicative of an event.^37^ In informal reviews of some participants’ medical histories, we found the same code referring to ectopic pregnancy or miscarriage repeated for years with no other indication of pregnancy, suggesting that in some cases these codes are carried forward in the problem list without representing new events. Future research on pregnancies that do not end in live birth will involve more thorough review to assess the accuracy of the algorithm for these outcomes. Furthermore, we followed Jones et al.^15^ in not attempting to distinguish spontaneous from induced abortion, which brings additional challenges.

## CONCLUSION

This pregnancy algorithm can be used by the community of researchers working on All of Us to identify pregnancy episodes and ask novel questions about experiences longitudinally with fertility, pregnancy, birth, and the postpartum and long-term health of pregnant people.

## Data Availability

All data used in the present study are freely available for registered researchers from the All of Us Research Program.

https://github.com/louisahsmith/allofus-pregnancy

## ACKNOWLEDGMENTS

We gratefully acknowledge *All of Us* participants for their contributions, without whom this and future pregnancy-related research would not be possible. We also thank the National Institutes of Health’s *All of Us* Research Program for making available the data examined in this study.

## SUPPLEMENTARY MATERIAL

**Supplementary Table 1.** Precision of the high-quality identified pregnancy episodes with respect to gestational age.

| **Precision category** | **n** | **percent** |
| --- | --- | --- |
| week | 10517 | 0.554 |
| week (poor support) | 2058 | 0.108 |
| two-week | 3185 | 0.168 |
| three-week | 1083 | 0.057 |
| month | 499 | 0.026 |
| two-month | 375 | 0.02 |
| three-month | 526 | 0.028 |
| non-specific | 727 | 0.038 |

**Supplementary Table 2.** Comparison of EHR data among pregnancies that were identified and confirmed by a positive survey response, compared to survey responses indicating pregnancy that were not linked to an identified episode.

|  | **Correctly**<br>**identified**<br>N = 3,832<br>mean(sd) | **Missed**<br>N = 848<br>mean(sd) | **p-value** |
| --- | --- | --- | --- |
| Total EHR codes | 90.1 (72.8) | 35.9 (69.2) | <0.001 |
| Total HIP codes | 12.0 (7.7) | 2.3 (5.7) | <0.001 |
| Abortion codes | 0.0 (0.2) | 0.0 (0.2) | 0.2 |
| Delivery codes | 2.1 (1.5) | 0.4 (0.9) | <0.001 |
| Ectopic pregnancy codes | 0.0 (0.3) | 0.0 (0.3) | 0.2 |
| Live birth codes | 1.4 (0.9) | 0.2 (0.6) | <0.001 |
| Pregnancy-only codes | 8.4 (6.3) | 1.5 (4.4) | <0.001 |
| Spontaneous abortion codes | 0.1 (0.6) | 0.1 (0.7) | 0.2 |
| Stillbirth codes | 0.0 (0.1) | 0.0 (0.1) | >0.9 |
| Mean days from survey (any code) | 62 (65) | -7 (85) | <0.001 |
| Mean days from survey (HIP code) | 80 (67) | 11 (99) | <0.001 |

**Supplementary Table 3.** Codes possibly related to pregnancy but not used in the pregnancy episode identification algorithm. Counts of codes across the All of Us dataset (after 2016) are compared to counts falling within the appropriate gestational timing of an identified pregnancy episode.

| **Concept** | **Total** | **Overlapping** | **Percent** |
| --- | --- | --- | --- |
| Breech presentation | 2297 | 1444 | 0.6286461 |
| Complication occurring during labor and delivery | 7172 | 4088 | 0.5699944 |
| Doppler velocimetry, fetal; umbilical artery | 2882 | 1956 | 0.6786954 |
| Excessive fetal growth affecting management of mother | 2107 | 1348 | 0.6397722 |
| False labor | 544 | 378 | 0.6948529 |
| False labor before 37 completed weeks of gestation | 1718 | 1376 | 0.8009313 |
| Fetal biophysical profile; with non-stress testing | 3151 | 2384 | 0.7565852 |
| Fetal biophysical profile; without non-stress testing | 13059 | 9998 | 0.7656023 |
| Fetal non-stress test | 13688 | 9543 | 0.6971800 |
| Gestation less than 24 weeks | 2043 | 1542 | 0.7547724 |
| Gestational diabetes mellitus | 11415 | 6909 | 0.6052562 |
| Initial prenatal care visit (report at first prenatal encounter with health care professional providing obstetrical care. Report also date of visit and, in a separate field, the date of the last menstrual period [LMP]) (Prenatal) | 1065 | 925 | 0.8685446 |
| Low lying placenta | 1026 | 821 | 0.8001949 |
| Medical genetics and genetic counseling services, each 30 minutes face-to-face with patient/family | 3231 | 909 | 0.2813370 |
| Polyhydramnios | 1534 | 1006 | 0.6558018 |
| Poor fetal growth affecting management | 4630 | 3260 | 0.7041037 |
| Pregnancy-induced hypertension | 3557 | 1910 | 0.5369694 |
| Prenatal flow sheet documented in medical record by first prenatal visit (documentation includes at minimum blood pressure, weight, urine protein, uterine size, fetal heart tones, and estimated date of delivery). Report also: date of visit and, in a separ | 210 | 158 | 0.7523810 |
| Preterm labor without delivery | 1268 | 896 | 0.7066246 |
| Reduced fetal movement | 2206 | 1635 | 0.7411605 |
| Requires diphtheria, tetanus and pertussis vaccination | 1979 | 260 | 0.1313795 |
| Suspected fetal abnormality affecting management of mother | 6329 | 4767 | 0.7531996 |
| Third trimester pregnancy | 53789 | 32378 | 0.6019446 |
| Ultrasound, pregnant uterus, real time with image documentation, fetal and maternal evaluation, after first trimester (> or = 14 weeks 0 days), transabdominal approach; single or first gestation | 6453 | 3983 | 0.6172323 |
| Ultrasound, pregnant uterus, real time with image documentation, transvaginal | 17651 | 11318 | 0.6412101 |

**Supplementary Table 4.** Probability ratios and 95% confidence intervals from a log-linear regression for the probability of multiple identified pregnancy episodes, among All of Us participants with at least one identified episode.

| Characteristic | Risk Ratio | 95% CI | p-value |
| --- | --- | --- | --- |
| **Race/ethnicity** |  |  |  |
| White | — | — |  |
| Asian | 1.03 | 0.83, 1.26 | 0.7 |
| Black or African-American | 1.23 | 1.08, 1.39 | <0.001 |
| Hispanic or Latino | 1.07 | 0.97, 1.19 | 0.11 |
| Middle Eastern or North African | 0.68 | 0.36, 1.15 | 0.14 |
| More than one race | 1.13 | 0.88, 1.41 | 0.2 |
| Native Hawaiian or Other Pacific Islander | 1.22 | 0.38, 2.86 | 0.6 |
| Other | 1.14 | 0.76, 1.62 | 0.4 |
| **Education** |  |  |  |
| College graduate | — | — |  |
| Less than high school | 1.01 | 0.84, 1.21 | 0.9 |
| High school graduate | 0.93 | 0.81, 1.08 | 0.3 |
| Some college | 0.90 | 0.79, 1.01 | 0.038 |
| Advanced degree | 1.15 | 1.02, 1.30 | 0.007 |
| **Family income** |  |  |  |
| > 100k | — | — |  |
| < 10k | 0.77 | 0.65, 0.91 | <0.001 |
| 10-25k | 0.77 | 0.65, 0.91 | <0.001 |
| 25-50k | 0.76 | 0.66, 0.88 | <0.001 |
| 50-100k | 0.79 | 0.70, 0.89 | <0.001 |
| **Marital status** |  |  |  |
| Married/partnered | — | — |  |
| Divorced/separated/widowed | 0.85 | 0.70, 1.01 | 0.038 |
| Never married | 0.75 | 0.67, 0.84 | <0.001 |
| **Age at first identified pregnancy episode** |  |  |  |
| 25-29 years | — | — |  |
| 15-19 years | 1.83 | 1.51, 2.20 | <0.001 |
| 20-24 years | 1.37 | 1.22, 1.53 | <0.001 |
| 30-34 years | 0.83 | 0.75, 0.92 | <0.001 |
| 35-39 years | 0.53 | 0.46, 0.61 | <0.001 |
| 40-44 years | 0.41 | 0.32, 0.53 | <0.001 |
| 45-49 years | 0.12 | 0.02, 0.37 | 0.002 |
| 50 years and over | 0.54 | 0.13, 1.40 | 0.3 |

**Supplementary Table 5.** Probability ratios and 95% confidence intervals from a log-linear regression for the probability of having a live birth, among all identified pregnancy episodes.

| Characteristic | Risk Ratio | 95% CI | p-value |
| --- | --- | --- | --- |
| **Race/ethnicity** |  |  |  |
| White | — | — |  |
| Asian | 1.08 | 0.95, 1.22 | 0.065 |
| Black or African-American | 0.97 | 0.89, 1.05 | 0.2 |
| Hispanic or Latino | 1.13 | 1.06, 1.20 | <0.001 |
| Middle Eastern or North African | 1.16 | 0.87, 1.51 | 0.084 |
| More than one race | 1.09 | 0.94, 1.26 | 0.058 |
| Native Hawaiian or Other Pacific Islander | 1.18 | 0.65, 1.96 | 0.3 |
| Other | 0.93 | 0.72, 1.18 | 0.4 |
| **Education** |  |  |  |
| College graduate | — | — |  |
| Less than high school | 0.96 | 0.86, 1.07 | 0.3 |
| High school graduate | 1.04 | 0.96, 1.13 | 0.13 |
| Some college | 1.02 | 0.95, 1.10 | 0.4 |
| Advanced degree | 0.98 | 0.91, 1.06 | 0.5 |
| **Family income** |  |  |  |
| > 100k | — | — |  |
| < 10k | 0.93 | 0.84, 1.03 | 0.036 |
| 10-25k | 1.00 | 0.91, 1.11 | >0.9 |
| 25-50k | 1.07 | 0.98, 1.17 | 0.015 |
| 50-100k | 1.06 | 0.98, 1.14 | 0.021 |
| **Marital status** |  |  |  |
| Married/partnered | — | — |  |
| Divorced/separated/widowed | 0.87 | 0.78, 0.96 | <0.001 |
| Never married | 0.90 | 0.85, 0.96 | <0.001 |
| **Age** |  |  |  |
| 25-29 years | — | — |  |
| 15-19 years | 0.99 | 0.84, 1.15 | 0.8 |
| 20-24 years | 1.07 | 0.99, 1.15 | 0.005 |
| 30-34 years | 1.0 | 0.93, 1.06 | 0.8 |
| 35-39 years | 0.93 | 0.87, 1.00 | 0.005 |
| 40-44 years | 0.83 | 0.73, 0.94 | <0.001 |
| 45-49 years | 0.63 | 0.41, 0.91 | 0.004 |
| 50 years and over | 0.16 | 0.03, 0.48 | 0.006 |

**Supplementary Table 6.** Probability ratios and 95% confidence intervals from a log-linear regression for the probability of preterm delivery (according to inferred gestational age), among all identified live births.

| Characteristic | Risk Ratio | 95% CI | p-value |
| --- | --- | --- | --- |
| **Race/ethnicity** |  |  |  |
| White | — | — |  |
| Asian | 0.86 | 0.61, 1.18 | 0.3 |
| Black or African-American | 1.18 | 1.01, 1.39 | 0.021 |
| Hispanic or Latino | 0.96 | 0.84, 1.09 | 0.5 |
| Middle Eastern or North African | 0.68 | 0.27, 1.38 | 0.3 |
| More than one race | 0.97 | 0.69, 1.32 | 0.8 |
| Native Hawaiian or Other Pacific Islander | 0.77 | 0.13, 2.39 | 0.7 |
| Other | 0.89 | 0.49, 1.48 | 0.6 |
| **Education** |  |  |  |
| College graduate | — | — |  |
| Less than high school | 1.77 | 1.40, 2.22 | <0.001 |
| High school graduate | 1.41 | 1.16, 1.71 | <0.001 |
| Some college | 1.59 | 1.34, 1.90 | <0.001 |
| Advanced degree | 1.08 | 0.89, 1.31 | 0.4 |
| **Family income** |  |  |  |
| > 100k | — | — |  |
| < 10k | 1.68 | 1.33, 2.12 | <0.001 |
| 10-25k | 1.47 | 1.17, 1.86 | <0.001 |
| 25-50k | 1.55 | 1.26, 1.91 | <0.001 |
| 50-100k | 1.34 | 1.10, 1.62 | 0.001 |
| **Marital status** |  |  |  |
| Married/partnered | — | — |  |
| Divorced/separated/widowed | 1.26 | 1.03, 1.52 | 0.005 |
| Never married | 0.92 | 0.81, 1.06 | 0.2 |
| **Age** |  |  |  |
| 25-29 years | — | — |  |
| 15-19 years | 1.20 | 0.87, 1.61 | 0.2 |
| 20-24 years | 0.95 | 0.81, 1.11 | 0.5 |
| 30-34 years | 1.13 | 0.98, 1.30 | 0.066 |
| 35-39 years | 1.42 | 1.21, 1.67 | <0.001 |
| 40-44 years | 1.38 | 1.05, 1.78 | 0.005 |
| 45-49 years | 3.05 | 1.57, 5.32 | <0.001 |
| 50 years and over | 4.74 | 0.27, 21.2 | 0.018 |

## Notes

### Competing Interest Statement

The authors have declared no competing interest.

### Funding Statement

This study did not receive any specific funding.

### Author Declarations

IRB of Northeastern University waived ethical approval for this work.

## REFERENCES

1. Shulman HB, D’Angelo DV, Harrison L, Smith RA, Warner L. The Pregnancy Risk Assessment Monitoring System (PRAMS): Overview of Design and Methodology. Am J Public Health. 2018;108(10):1305–1313. doi:10.2105/AJPH.2018.304563

2. Schoendorf KC, Branum AM. The use of United States vital statistics in perinatal and obstetric research. Am J Obstet Gynecol. 2006;194(4):911–915. doi:10.1016/j.ajog.2005.11.020

3. Kishi R, Kobayashi S, Ikeno T, et al. Ten years of progress in the Hokkaido birth cohort study on environment and children’s health: cohort profile—updated 2013. Environ Health Prev Med. 2013;18(6):429–450. doi:10.1007/s12199-013-0357-3

4. Magnus P, Irgens LM, Haug K, Nystad W, Skjærven R, Stoltenberg C. Cohort profile: The Norwegian Mother and Child Cohort Study (MoBa). Int J Epidemiol. 2006;35(5):1146–1150. doi:10.1093/ije/dyl170

5. Niswander KR, Gordon MJ, Gordon M. The Women and Their Pregnancies: The Collaborative Perinatal Study of the National Institute of Neurological Diseases and Stroke. National Institute of Health; 1972.

6. Wise LA, Rothman KJ, Mikkelsen EM, et al. Design and conduct of an internet-based preconception cohort study in north america: pregnancy study online. Paediatr Perinat Epidemiol. 2015;29(4):360–371. doi:10.1111/ppe.12201

7. Voorst SF van, Vos AA, Jong-Potjer LC de, Waelput AJM, Steegers EAP, Denktaş S. Effectiveness of general preconception care accompanied by a recruitment approach: protocol of a community-based cohort study (the Healthy Pregnancy 4 All study). BMJ Open. 2015;5(3):e006284. doi:10.1136/bmjopen-2014-006284

8. Spry E, Olsson CA, Hearps SJC, et al. The Victorian Intergenerational Health Cohort Study (VIHCS): Study design of a preconception cohort from parent adolescence to offspring childhood. Paediatr Perinat Epidemiol. 2020;34(1):86–98. doi:10.1111/ppe.12602

9. Loo EXL, Soh SE, Loy SL, et al. Cohort profile: Singapore Preconception Study of Long-Term Maternal and Child Outcomes (S-PRESTO). Eur J Epidemiol. 2021;36(1):129–142. doi:10.1007/s10654-020-00697-2

10. Daw JR, Auty SG, Admon LK, Gordon SH. Using Modernized Medicaid Data to Advance Evidence-Based Improvements in Maternal Health. Am J Public Health. 2023;113(7):805–810. doi:10.2105/AJPH.2023.307287

11. The “All of Us” research program. N Engl J Med. 2019;381(7):668–676. doi:10.1056/nejmsr1809937

12. Gomez SE, Sarraju A, Rodriguez F. Racial and Ethnic Group Underrepresentation in Studies of Adverse Pregnancy Outcomes and Cardiovascular Risk. J Am Heart Assoc. 2022;11(5):e024776. doi:10.1161/JAHA.121.024776

13. Girardi G, Longo M, Bremer AA. Social determinants of health in pregnant individuals from underrepresented, understudied, and underreported populations in the United States. Int J Equity Health. 2023;22(1):186. doi:10.1186/s12939-023-01963-x

14. Moseson H, Fix L, Hastings J, et al. Pregnancy intentions and outcomes among transgender, nonbinary, and gender-expansive people assigned female or intersex at birth in the United States: Results from a national, quantitative survey. Int J Transgender Health. 2020;22(1-2):30–41. doi:10.1080/26895269.2020.1841058

15. Jones SE, Bradwell KR, Chan LE, et al. Who is pregnant? Defining real-world data-based pregnancy episodes in the National COVID Cohort Collaborative (N3C). JAMIA Open. 2023;6(3):ooad067. doi:10.1093/jamiaopen/ooad067

16. Stang PE, Ryan PB, Racoosin JA, et al. Advancing the science for active surveillance: rationale and design for the observational medical outcomes partnership. Ann Intern Med. 2010;153(9):600. doi:10.7326/0003-4819-153-9-201011020-00010

17. Observational Health Data Sciences and Informatics. The Book of OHDSI.; 2021. Accessed April 4, 2024. https://ohdsi.github.io/TheBookOfOhdsi/

18. Wickham H, Girlich M, Ruiz E. Dbplyr: A “dplyr” Back End for Databases.; 2024. https://CRAN.R-project.org/package=dbplyr

19. Smith LH, Cavanaugh R. allofus R package. Published online December 21, 2023. doi:10.5281/ZENODO.10420610

20. Centers for Disease Control and Prevention, National Center for Health Statistics. National vital statistics system, natality. Accessed November 27, 2023. http://wonder.cdc.gov/natality-expanded-current.html

21. Fleszar LG, Bryant AS, Johnson CO, et al. Trends in State-Level Maternal Mortality by Racial and Ethnic Group in the United States. JAMA. 2023;330(1):52–61. doi:10.1001/jama.2023.9043

22. Hoyert D. Maternal Mortality Rates in the United States, 2021. National Center for Health Statistics (U.S.); 2023. doi:10.15620/cdc:124678

23. Romero R, Dey SK, Fisher SJ. Preterm labor: One syndrome, many causes. Science. 2014;345(6198):760–765. doi:10.1126/science.1251816

24. Mitrogiannis I, Evangelou E, Efthymiou A, et al. Risk factors for preterm birth: an umbrella review of meta-analyses of observational studies. BMC Med. 2023;21(1):494. doi:10.1186/s12916-023-03171-4

25. Data Sources – All of Us Research Hub. Accessed April 4, 2024. https://www.researchallofus.org/data-tools/data-sources/

26. Modde Epstein C, McCoy TP. Linking Electronic Health Records With Wearable Technology From the All of Us Research Program. J Obstet Gynecol Neonatal Nurs. 2023;52(2):139–149. doi:10.1016/j.jogn.2022.12.003

27. Yhdego HH, Nayebnazar A, Amrollahi F, et al. Prediction of Unplanned Hospital Readmission using Clinical and Longitudinal Wearable Sensor Features. Published online April 11, 2023:2023.04.10.23288371. doi:10.1101/2023.04.10.23288371

28. Ramirez AH, Sulieman L, Schlueter DJ, et al. The All of Us Research Program: Data quality, utility, and diversity. Patterns. 2022;3(8):100570. doi:10.1016/j.patter.2022.100570

29. Canelón SP, Burris HH, Levine LD, Boland MR. Development and evaluation of MADDIE: method to acquire delivery date information from electronic health records. Int J Med Inf. 2021;145:104339. doi:10.1016/j.ijmedinf.2020.104339

30. Chomistek AK, Phiri K, Doherty MC, et al. Development and Validation of ICD-10-CM-based Algorithms for Date of Last Menstrual Period, Pregnancy Outcomes, and Infant Outcomes. Drug Saf. 2023;46(2):209–222. doi:10.1007/s40264-022-01261-5

31. Zhu Y, Bateman BT, Hernandez-Diaz S, et al. Validation of claims-based algorithms to identify non-live birth outcomes. Pharmacoepidemiol Drug Saf. 2023;32(4):468–474. doi:10.1002/pds.5574

32. Devine S, West S, Andrews E, et al. The identification of pregnancies within the general practice research database. Pharmacoepidemiol Drug Saf. 2010;19(1):45–50. doi:10.1002/pds.1862

33. Li Q, Andrade SE, Cooper WO, et al. Validation of an algorithm to estimate gestational age in electronic health plan databases. Pharmacoepidemiol Drug Saf. 2013;22(5):524–532. doi:10.1002/pds.3407

34. Matcho A, Ryan P, Fife D, Gifkins D, Knoll C, Friedman A. Inferring pregnancy episodes and outcomes within a network of observational databases. PLOS ONE. 2018;13(2):e0192033. doi:10.1371/journal.pone.0192033

35. Sarayani A, Wang X, Thai TN, Albogami Y, Jeon N, Winterstein AG. Impact of the Transition from ICD–9–CM to ICD–10–CM on the Identification of Pregnancy Episodes in US Health Insurance Claims Data. Clin Epidemiol. 2020;Volume 12:1129–1138. doi:10.2147/CLEP.S269400

36. Ailes EC, Zhu W, Clark EA, et al. Identification of pregnancies and their outcomes in healthcare claims data, 2008–2019: An algorithm. PLOS ONE. 2023;18(4):e0284893. doi:10.1371/journal.pone.0284893

37. Scholes D, Yu O, Raebel MA, Trabert B, Holt VL. Improving automated case finding for ectopic pregnancy using a classification algorithm. Hum Reprod. 2011;26(11):3163–3168. doi:10.1093/humrep/der299

